# The levels distribution of the serum lipids in Xinjiang adults, 2018

**DOI:** 10.1101/2024.12.12.24318932

**Authors:** Adila Sulidan, Xiamusiye Muyiduli, Jun Zhang, Chunmei Ling, Maidina Abudusilimu, Parman Mardan, Yilixiati Kulaixi, Siyu Zhang, Yongqi Ding, Rong Zhang

## Abstract

**Objective:** To analyze the prevalence and distribution characteristics of dyslipidemia in adult residents of Xinjiang Uygur Autonomous Region (Xinjiang), and to provide effective prevention and control measures for dyslipidemia.

**Methods:** In 2018, a total of 4865 permanent residents aged 18 years and above were investigated by multi-stage stratified cluster random sampling in 8 monitoring counties (cities and districts) in Xinjiang. Questionnaire survey and laboratory tests were conducted, and fasting venous blood was collected to detect the levels of TC, TG, HDL-C and LDL-C in serum. To analyze the difference of the detection results of relevant indexes in serum of different age, sex and urban and rural adults.

**Results:** The prevalence of dyslipidemia in adults in Xinjiang was 48.35%, among which the prevalence of hypercholesterolemia(high TC), hypertriglyceridemia(high TG), high low-density lipoprotein cholesterol(high LDL-C) and low high-density lipoprotein cholesterol (low HDL-C)were 18.58%, 28.04%, 25.30% and 13.40%. In 2018, the serum TC level in Xinjiang adults was (4.46±0.91) mmol/L, and that in males and females was (4.42±0.89) and (4.49±0.92) mmol/L, respectively (*t*=-2.848, *p* < 0.05). The serum TC level of urban residents (4.57±0.93) mmol/L was higher than that of rural residents (4.38±0.89) mmol/L, and the difference was statistically significant (*t*=6.979, *p* < 0.001). The difference of serum TC levels in different age groups was statistically significant (*F*=199.389, *p* < 0.001). The serum TG level was (1.52±1.12) mmol/L, (1.64±1.31) in male and (1.43±0.93) mmol/L in female, (*t*=6.359, *p* < 0.001). The serum TG level of urban residents (1.66±1.33) mmol/L was higher than that of rural residents (1.43±0.94) mmol/L, and the difference was statistically significant (*t*=7.088, *p* < 0.001). There were significant differences in serum TG levels among different age groups (*F*=31.355, *p* < 0.001). Serum HDL-C level was (1.23±0.33) mmol/L, (1.14±0.32) in male and (1.29±0.33) mmol/L in female, (*t*=-16.033, *p* < 0.001). The serum HDL-C level of urban residents (1.25±0.34 mmol/L) was higher than that of rural residents (1.21±0.33 mmol/L), and the difference was statistically significant (*t*=4.298, *p* < 0.001). The difference of serum HDL-C level in different age groups was statistically significant (*F*=10.992, *p* < 0.001). Serum LDL-C level was (2.55±0.77) mmol/L, (2.54±0.76) in male and (2.55±0.78) mmol/L in female, (*t*=-0.426, *p* > 0.05). The serum LDL-C level of urban residents (2.58±0.82) mmol/L was higher than that of rural residents (2.53±0.74) mmol/L, and the difference was statistically significant (*t*=2.180, *p* < 0.05). There were significant differences in serum LDL-C levels among different age groups (*F*=121.679, *p* < 0.001).

**Conclusion:** The prevalence of dyslipidemia in adults in Xinjiang is higher than the national level, the serum LDL-C level in men is lower than that in women, and the serum TC, TG and HDL-C levels are higher than that in women.

## 1 Introduction

With the continuous improvement of China’s economic and social development and health service level, the average life expectancy of residents continues to increase. With the continuous extension of the survival period of patients with chronic diseases, coupled with the aging of population, the acceleration of urbanization, industrialization and the prevalence of behavioral risks, the incidence of cardiovascular and cerebrovascular diseases has risen, becoming a prominent problem threatening the health of residents. Dyslipidemia is a basic factor in the formation of atherosclerosis, plays an important role in its formation and development, and is an important risk factor for cardiovascular diseases [1-2].

According to the Report on Nutrition and Chronic Diseases of Chinese Residents 2020 [3], the total prevalence rate of dyslipidemia in adults ≥ 18 years old in China was 35.6%, an increase of 17% compared with 2002 [4]. Therefore, effective control of dyslipidemia is of great significance for the prevention and control of cardiovascular and cerebrovascular diseases. In order to understand the distribution characteristics of blood lipid levels in adults in Xinjiang, the relevant investigation results of chronic diseases and their risk factors monitoring in Chinese adults in Xinjiang in 2018 were studied.

## 2 Objects and methods

### 2.1 Research object

In July to November 2018, the monitoring project of chronic diseases and risk Factors in Chinese adults in Xinjiang covered 8 counties (cities and districts), and the survey objects were local permanent residents aged 18 years and above (who had lived in the survey place for more than 6 months), and finally included 4865 subjects for analysis°The project was reviewed by the Ethics Review Committee of the Chinese Center for Disease Control and Prevention (CDC), and all investigation subjects signed informed consent.

### 2.2 Survey Methods

A multi-stage stratified cluster random sampling method is adopted, and a systematic sampling based on population size ranking is adopted to randomly select 3 townships (subdistricts and regiments). In each selected township (subdistricts and regiments), 2 administrative villages (neighborhood committees and companies) are randomly selected. In each selected administrative village (neighborhood committees and companies), At least 60 households were divided into several villager/resident groups, and one villager/resident group was selected by simple random sampling method. In the fourth stage of sampling, 45 households were selected from each selected villager/resident group, and questionnaires and laboratory tests were conducted on chronic diseases and nutrition of permanent residents aged 18 and above in the survey households [5].

### 2.3 Questionnaire

(1) Adopt a nationally designed questionnaire and conduct the survey by uniformly trained investigators,The content includes general information such as gender, age, and place of residence.(2) Laboratory examination The fasting venous blood of the subjects was collected, and total cholesterol (TC), triglyceride(TG),high-density lipoprotein cholesterol(HDL-C),low density lipoprotein cholesterol (LDL-C),were detected by a third-party testing company using an automatic biochemical analyzer.

### 2.4 Classification and diagnostic criteria

According to the Chinese Guideline for the Management of Dyslipidemia in Adults (2016) [6], High TC was defined as TC ≥ 6.22 mmol/L,High LDL-C was defined as LDL-C ≥ 4.14 mmol/L, Low HDL-C was defined as HDL-C< 1.04 mmol/L and high TG was defined as TG ≥ 2.26 mmol/L. and abnormal appearance of any of the above indicators is judged as dyslipidemia. Those who meet any of the above conditions or are diagnosed as dyslipidemia by community health service centers/township health centers or superior medical and health institutions are dyslipidemia.

### 2.5 Quality Control

Investigators are trained at the national level (level 1) or provincial level (Level 2) and participate in the investigation work after passing the assessment.Quality control samples and blind samples are provided by the national laboratory, and 8 monitoring points pass the blind sample assessment before starting field work.For every 30 samples measured, a pair of fixed value quality control was made for quality control. The collection, storage, transportation and whole quality control of blood samples were conducted. The blood samples were stored at −70°C away from light.

### 2.6 Statistical Analysis

SPSS26.0 software was used to describe and analyze the data statistically. The demographic distribution was described by the composition ratio, and the detection rate of dyslipidemia was described by the χ2 test. The mean value between genders and regions was compared by the T-test, and the mean value between ages was compared by the analysis of variance. *P* < 0.05 was considered statistically significant.

## 3 Results

## 3.1 Basic Information

A total of 4865 subjects were surveyed, among which 45-59 years old had the highest proportion and 60 years old ≥ had the lowest proportion. Women are higher than men; More rural than urban areas (Table 1).

**Table 1.** Gender distribution of respondents with different characteristics (n(%))

| characteristic | male | female | Both |
| --- | --- | --- | --- |
| Age (years old) |  |  |  |
| 18~44 | 752 (34.54) | 1013 (37.69) | 1765 (36.28) |
| 45~59 | 774 (35.55) | 1051 (39.10) | 1825 (37.51) |
| ≥60 | 651 (29.90) | 624 (23.21) | 1275 (26.21) |
| Urban and rural |  |  |  |
| Urban area | 783 (35.95) | 1146 (42.63) | 1929 (39.64) |
| Rural area | 1395 (64.05) | 1542 (57.37) | 2937 (60.36) |
| Both | 2177 (44.75) | 2688 (55.25) | 4865 (100.00) |

### 3.2 Prevalence of dyslipidemia

Among 4865 adult residents, 2352 had dyslipidemia, and the prevalence of dyslipidemia was 48.35%, among which the prevalence of hypercholesterolemia(high TC), hypertriglyceridemia(high TG), high low-density lipoprotein cholesterol(high LDL-C) and low high-density lipoprotein cholesterol (low HDL-C)l were 18.58%, 28.04%, 25.30% and 13.40%, respectively.Different characteristics of residents compare,There were statistically significant differences in the prevalence of dyslipidemia, high TC, high TG and low HDL-C among different genders (P < 0.001 for all) (Table 2).

**Table 2.** Comparison of dyslipidemia among urban and rural residents with different characteristics.

|  |  | Dyslipemia |  | High TC |  | High TG |  | High LDL-C |  | Low HDL-C |  |
| --- | --- | --- | --- | --- | --- | --- | --- | --- | --- | --- | --- |
| characteristic |  | Number of cases | Prevalence rate | Number of cases | Prevalence rate | Number of cases | Prevalence rate | Number of cases | Prevalence rate | Number of cases | Prevalence rate |
| Gender | male | 1182 | 50.3 | 347 | 38.4 | 678 | 49.7 | 761 | 61.8 | 280 | 42.9 |
|  | female | 1170 | 49.7* | 557 | 61.6* | 686 | 50.3* | 470 | 38.2 | 372 | 57.1* |
| Age (years old) | 18-44 | 698 | 29.7 | 173 | 19.1 | 346 | 25.3 | 488 | 39.6 | 123 | 18.9 |
|  | 45-59 | 936 | 39.8 | 396 | 43.8 | 564 | 41.3 | 439 | 35.7 | 289 | 44.3 |
|  | ≥60 | 718 | 30.5* | 335 | 37.1* | 454 | 33.3* | 304 | 24.7* | 240 | 36.8* |
| Urban/Rural | Urban area | 1001 | 42.6 | 426 | 47.1 | 653 | 47.9 | 453 | 36.8 | 316 | 48.5 |
|  | Rural area | 1351 | 57.4* | 478 | 52.9* | 711 | 52.1* | 778 | 63.2* | 336 | 51.5* |

Intra-group comparison \* $p < 0.001$

### 3.3 Blood lipid and serum levels of residents

#### 3.3.1 Serum TC level

The serum TC level of Xinjiang adults was (4.46±0.91) mmol/L. Women were higher than men, and the difference between men and women was statistically significant (*t*=-2.848, *p* < 0.05).The highest value was (4.52±0.84) mmol/L in males aged 45-59 years and (4.92±0.92) mmol/L in females aged ≥60 years.The serum TC level of urban residents was higher than that of rural residents, and the difference was statistically significant (*t*=6.979, *p* < 0.001).The difference of serum TC levels in different age groups was statistically significant (*F*=199.389, *p* < 0.001). The level of serum TC in blood lipid increased with age (Table 3).

**Table 3.** TC levels of residents in different regions 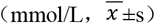.

| Gender | Urban area | Rural area | Both |
| --- | --- | --- | --- |

| Age (years old) |  |  |  |
| --- | --- | --- | --- |
| male |  |  |  |
| 18-44 | 4.53±0.99 | 4.18±0.90 | 4.26±0.93 |
| 45-59 | 4.55±0.92 | 4.51±0.80 | 4.52±0.84 |
| ≥60 | 4.42±0.91 | 4.53±0.82 | 4.48±0.87 |
| Total | 4.49±0.93 | 4.38±0.86 | 4.42±0.89 |
| female |  |  |  |
| 18-44 | 4.05±0.75 | 4.02±0.78 | 4.03±0.77 |
| 45-59 | 4.73±0.84 | 4.64±0.89 | 4.68±0.87 |
| ≥60 | 4.95±0.95 | 4.89±0.88 | 4.92±0.92 |
| Total | 4.63±0.92 | 4.39±0.91 | 4.49±0.92 |
| Both |  |  |  |
| 18-44 | 4.22±0.87 | 4.09±0.84 | 4.13±0.85 |
| 45-59 | 4.66±0.87 | 4.58±0.85 | 4.62±0.86 |
| ≥60 | 4.69±0.97 | 4.70±0.87 | 4.69±0.92 |
| Total | 4.57±0.93 | 4.38±0.89 | 4.46±0.91 |

Comparison of male levels in different age groups: $F=18.728$ , $p < 0.001$ ; Comparison of female levels in different age groups: $F=254.679$ , $p < 0.001$ ; Comparison of different age groups in Urban areas: $F=43.126$ , $p < 0.001$ ; Comparison of different age groups in rural areas: $F=140.022$ , $p < 0.001$ .

#### 3.3.2 Serum TG leve

The serum TG level of Xinjiang adults was (1.52±1.12) mmol/L. The difference between males and females was statistically significant (*t*=6.359, *p* < 0.001).The highest value was (1.68±1.53) mmol/L in males aged 18-44 years and (1.77±1.11) mmol/L in females aged ≥60 years.The serum TG level of urban residents was higher than that of rural residents, and the difference was statistically significant (*t*=7.088, *p* < 0.001).There were significant differences in serum TG levels among different age groups (*F*=31.355, *p* < 0.001). Blood lipid and serum TG levels of residents increased with age (Table 4).

**Table 4.** TG levels of residents in different regions 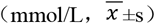.

| Gender |  |  |  |
| --- | --- | --- | --- |
|  | Urban area | Rural area | Both |
| Age (years old) |  |  |  |
| male |  |  |  |
| 18-44 | 1.92±2.21 | 1.61±1.28 | 1.68±1.53 |
| 45-59 | 1.96±1.84 | 1.51±0.83 | 1.68±1.32 |
| ≥60 | 1.64±0.95 | 1.44±0.99 | 1.55±0.98 |
| Total | 1.82±1.62 | 1.54±1.08 | 1.64±1.31 |
| female |  |  |  |
| 18-44 | 1.10±0.60 | 1.14±0.66 | 1.13±0.64 |
| 45-59 | 1.59±1.06 | 1.46±0.84 | 1.53±0.95 |
| ≥60 | 1.89±1.26 | 1.61±0.84 | 1.77±1.11 |
| Total | 1.56±1.08 | 1.34±0.78 | 1.43±0.93 |
| Both |  |  |  |
| 18-44 | 1.38±1.45 | 1.35±1.01 | 1.36±1.14 |
| 45-59 | 1.73±1.41 | 1.49±0.84 | 1.59±1.13 |
| ≥60 | 1.77±1.13 | 1.52±0.93 | 1.66±1.05 |
| Total | 1.66±1.33 | 1.43±0.94 | 1.52±1.12 |

Comparison of male levels in different age groups: $F=2.285$ , $p > 0.05$ ; Comparison of female levels in different age groups: $F=98.602$ , $p < 0.001$ ; Comparison of different age groups in Urban areas: $F=12.971$ , $p < 0.001$ ;Comparison of different age groups in rural areas: $F=9.068$ , $p < 0.001$ .

#### 3.3.3 Serum HDL-C level

The serum HDL-C level of adult residents in Xinjiang was (1.23±0.33) mmol/L.There was a statistically significant difference between males and females (*t*=-16.033, *p* < 0.001). Serum HDL-C level in males increased with age, and the highest level was (1.31±0.34) mmol/L in females aged 45-59 years.The serum HDL-C level of urban residents was higher than that of rural residents, and the difference was statistically significant (*t*=4.298, *p* < 0.001).The difference of serum HDL-C level in different age groups was statistically significant (*F*=10.992, *p* < 0.001). The level of serum HDL-C in residents aged 45 ∼59 years was the highest (Table 5).

**Table 5.**
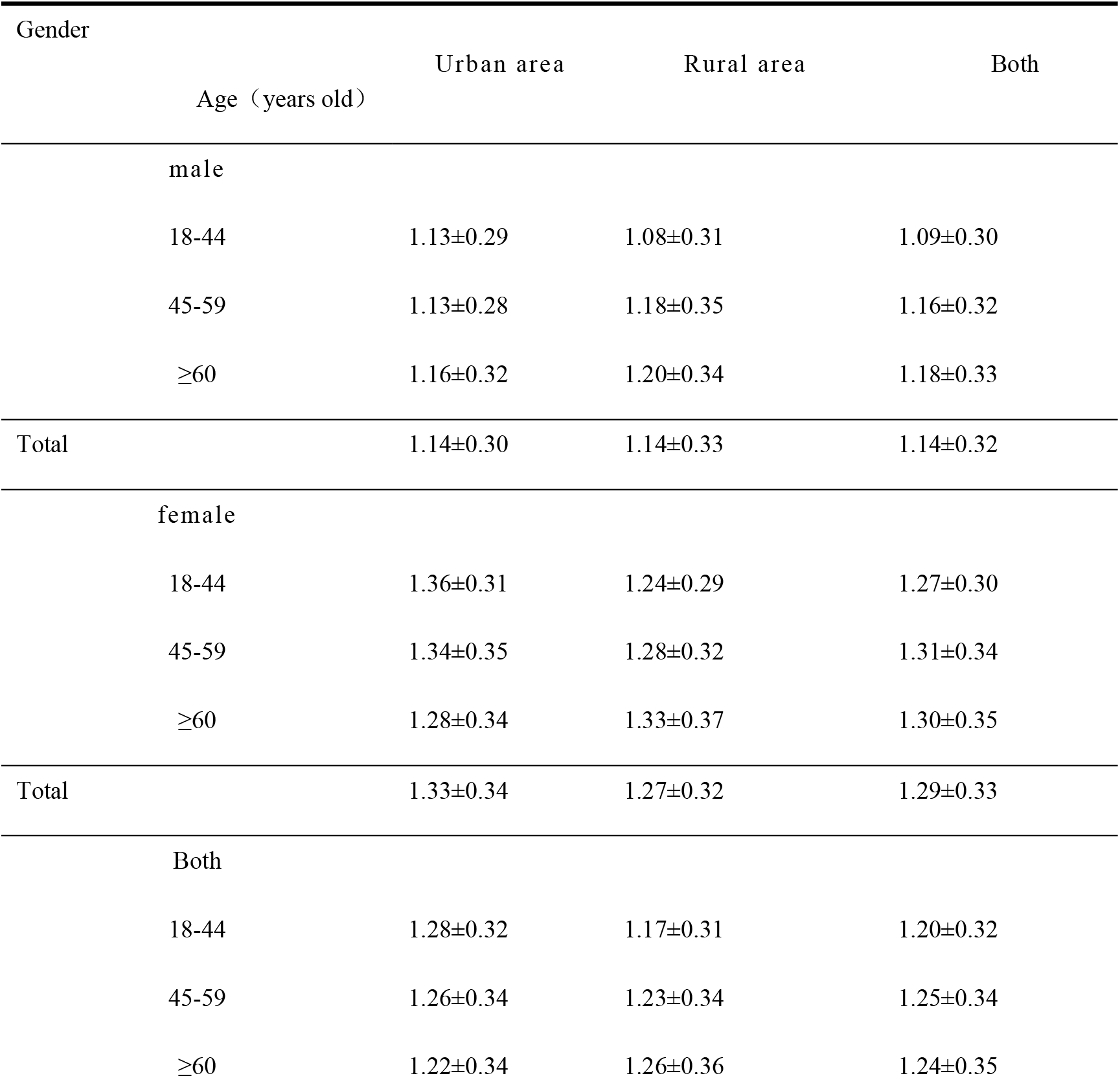
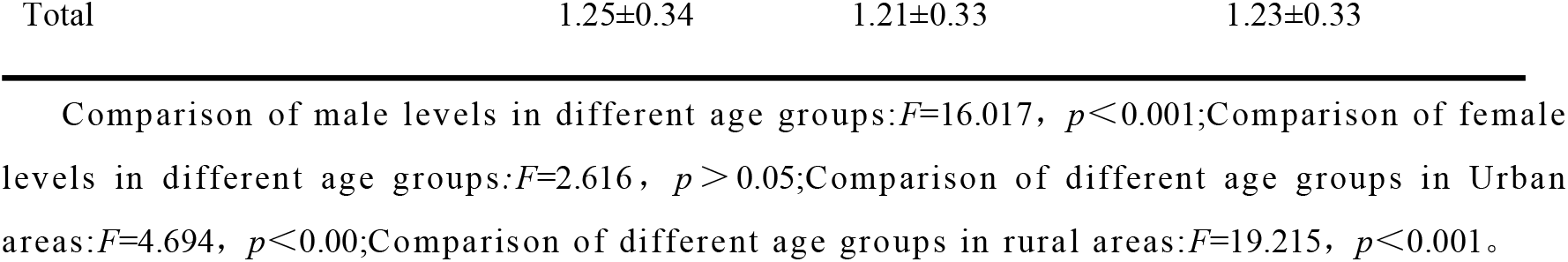
HDL-C levels of residents in different regions 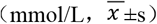.

#### 3.3.4 Serum LDL-C level

The serum LDL-C level of adult residents in Xinjiang was (2.55±0.77) mmol/L.There was no significant difference between males and females (*t*=-0.426, *p* > 0.05). The serum LDL-C level in males was the highest at 45 ∼ 59 years old ((2.61±0.73) mmol/L), and it increased with age in females.The serum LDL-C level of urban residents was higher than that of rural residents, and the difference was statistically significant (*t*=2.180, *p* < 0.05).There were significant differences in serum LDL-C levels among different age groups (*F*=121.679, *p* < 0.001). The level of serum LDL-C increased with age (Table 6).

**Table 6.**
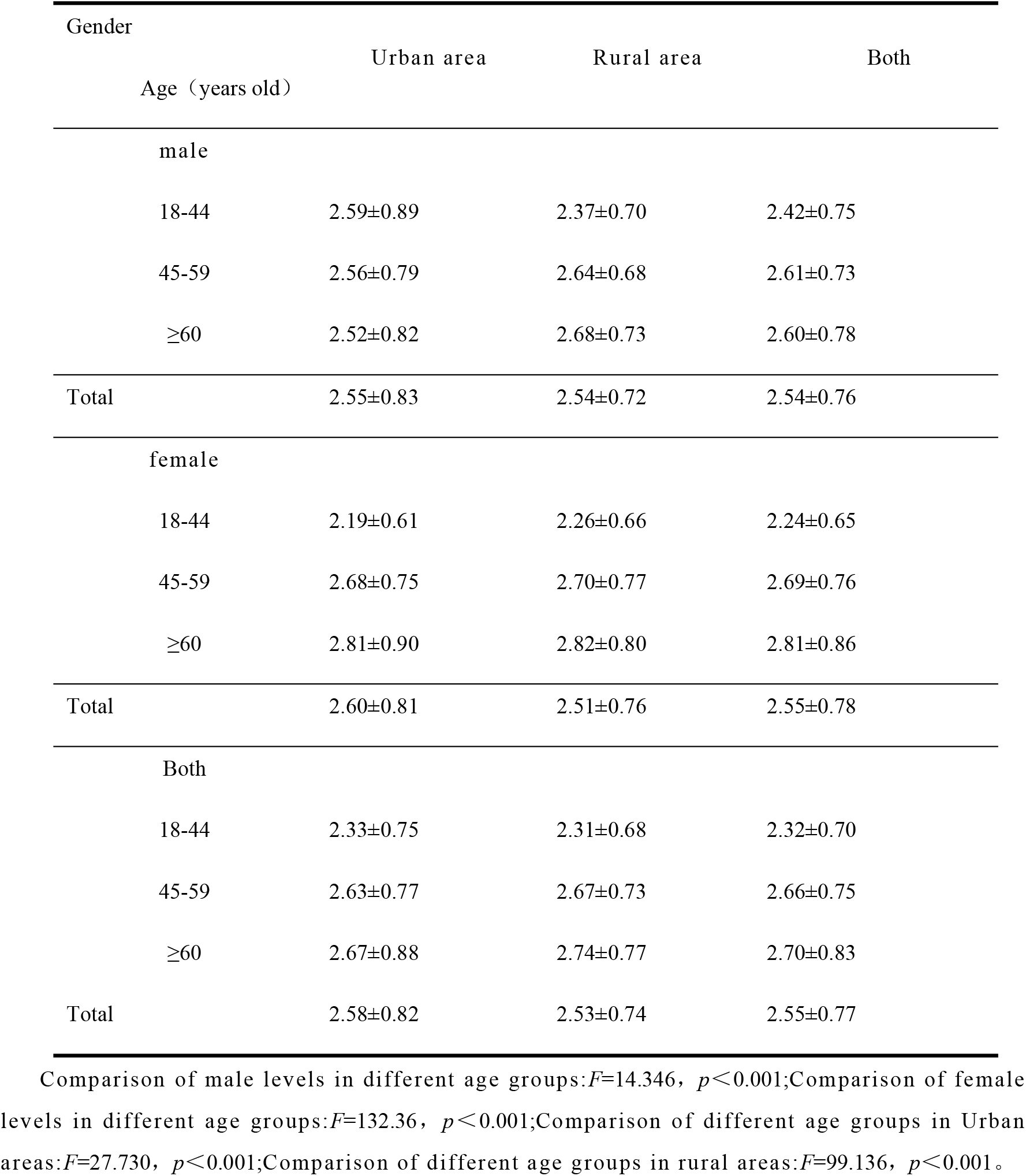
LDL-C levels of residents in different regions 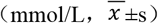.

## 4 Discussion

A large number of epidemiological studies have confirmed that dyslipidemia is one of the important risk factors for cardiovascular diseases. With the development of China’s economy, people’s lifestyle has changed greatly, and the incidence of hyperlipidemia and other diseases has increased significantly.Effective prevention and treatment of hyperlipidemia has great medical and social significance at present.

The results of this study showed that the prevalence of dyslipidemia in adult residents of Xinjiang was 48.35%, higher than that of adult residents of Xinjiang Uygur Autonomous Region from 2013 to 2014 (45.00%) [7].Compared with other regions, it was higher than that of China (35.6%)[3], Qingdao (40.53%)[8] Shanxi Province (31.70%)[9] lower than that of an urban area in Anhui Province (49.4%) [10] and Yan ‘an City (57.75%) [11].It may be related to the different economic development level, residents’ lifestyle, dietary structure, residents’ health knowledge and awareness and other factors.

In this study, it was found that hypertriglyceridemia(High TG) and high low-density lipoprotein cholesterol(High LDL-C) were the main types of dyslipidemia in urban and rural residents in Xinjiang, with prevalence rates of 28.04% and 25.30%, respectively,which was different from that in the whole country,hypertriglyceridemia(High TG) and low high-density lipoprotein cholesterol (Low HDL-C) were the main types of dyslipidemia[12]. Whether it is the geographical difference or the representativeness of the sample needs further study.

high TC, high TG and high LDL-C a were all higher than the national levels [12]. The prevalence of low HDL-C was 13.40%, which was significantly lower than that of the whole country (20.9%) [12] and Hainan Province (16.3%) [13].

The results showed that the prevalence of dyslipidemia and high LDL-C in men was higher than that in women, which may be related to the factors such as more work pressure, frequent social activities, unhealthy diet and lifestyle in men.The prevalence of high TC, high TG and low HDL-C in women was higher than that in men.The prevalence of dyslipidemia,high TC, high TG and low HDL-C in 45-59 years old were higher than those in other age groups.The prevalence of various dyslipidemia in rural areas is higher than that of urban residents, which is different from the results of Ma Teng et al [14] in Xinjiang. The accessibility of medical and public health services in rural residents is lower than that in urban areas, and the level of education is also lower.

The results of this study found that there were significant gender differences, age differences and urban and rural differences in blood lipid composition of Xinjiang residents. The serum levels of TC, LDL-C and HDL-C in males were higher than those in males.The levels of various lipid components of women over 45 years old are higher than those of women aged 18-44 years old, and higher than those of men in the same age group, indicating that the lipid levels of women during perimenopause and menopause increase significantly. Estrogen plays an important role in regulating lipid levels, and with the decrease of hormone levels, the lipid status is further affected [15].The levels of various lipid components in urban areas are higher than those in rural areas, indicating that with the rapid economic development, the improvement of people’s living standards and the change of lifestyle, the prevalence of dyslipidemia is on the rise rapidly [16].The lipid levels of the adult residents in Xinjiang were 4.46 mmol/L TC, 1.43 mmol/L TG, 2.55 mmol/L LDL-C and 1.23 mmol/L HDL-C. Lower than the national level (TC 4.70 mmol/L, TG 1.49 mmol/L, LDL-C 2.88 mmol/L, HDL-C 1.35 mmol/L) [17].

To sum up, the levels of various lipid components of urban and rural residents in Xinjiang are lower than the national level, but the prevalence of dyslipidemia is relatively high, and the prevention and control situation is severe. In particular, it is necessary to focus on the prevention and control of hypertriglyceridemia and high low-density lipoprotein cholesterol, focusing on middle-aged and elderly female residents, and formulating effective prevention and control measures according to the distribution characteristics of lipid levels in adults in Xinjiang.Reduce the prevalence of dyslipidemia and reduce the risk of cardiovascular and cerebrovascular diseases caused by dyslipidemia.

## 5 Conclusion

The prevalence of dyslipidemia in adults in Xinjiang was higher than the national level, the serum LDL-C level in men was lower than that in women, and the serum TC, TG and HDL-C levels were higher than that in women. Serum levels of TC, TG, LDL-C and HDL-C were different among different age groups and urban and rural residents°

## Data Availability

All relevant data are within the manuscript and its Supporting Information files. ?

